# Clinical characterization and Genomic analysis of COVID-19 breakthrough infections during second wave in different states of India

**DOI:** 10.1101/2021.07.13.21260273

**Authors:** Nivedita Gupta, Harmanmeet Kaur, Pragya Yadav, Labanya Mukhopadhyay, Rima R Sahay, Abhinendra Kumar, Dimpal A. Nyayanit, Anita M. Shete, Savita Patil, Triparna Majumdar, Salaj Rana, Swati Gupta, Jitender Narayan, Neetu Vijay, Pradip Barde, Gita Natrajan, B Amurtha Kumari, Manasa P Kumari, Debasis Biswas, Jyoti Iravane, Sharmila Raut, Shanta Dutta, Sulochana Devi, Purnima Barua, Piyali Gupta, Biswa Borkakoty, Deepjyoti Kalita, Kanwardeep Dhingra, Bashir Fomda, Yash Joshi, Kapil Goyal, Reena John, Ashok, Rahul Dhodapkar, Priyanka Pandit, Sarada Devi, Manisha Dudhmal, Deepa Kinariwala, Neeta Khandelwal, Yogendra Kumar Tiwari, PK Khatri, Anjali Gupta, Himanshu Khatri, Bharti Malhotra, Mythily Nagasundaram, Lalit Dar, Nazira Sheikh, Neeraj Aggarwal, Priya Abraham

**Author notes:** **Corresponding author:** Dr Pragya D Yadav, Scientist ‘E’ and Group Leader, Maximum Containment Facility, Indian Council of Medical Research- National Institute of Virology, Sus Road, Pashan, Pune, India Pin-411021.

## Abstract

During March to June 2021 India has experienced a deadly second wave of COVID-19 with an increased number of post-vaccination breakthrough infections reported across the country. To understand the possible reason of these breakthroughs we collected 677 clinical samples (throat swab/ nasal swabs) of individuals who had received two doses (n=592) and one dose (n=85) of vaccines (Covishield and Covaxin,) and tested positive for COVID-19, from 17 states/Union Territories of country. These cases were telephonically interviewed and clinical data was analyzed. A total of 511 SARS-CoV-2 genomes were recovered with genome coverage of higher than 98% from both the cases. Analysis of both the cases determined that 86.69% (n=443) of them belonged to the Delta variant along with Alpha, Kappa, Delta AY.1 and Delta AY.2. The Delta variant clustered into 4 distinct sub-lineages. Sub-lineage–I had mutations: ORF1ab-A1306S, P2046L, P2287S, V2930L, T3255I, T3446A, G5063S, P5401L, A6319V and N-G215C; Sub-lineage –II : ORF1ab- P309L, A3209V, V3718A, G5063S, P5401L and ORF7a-L116F; Sub-lineage –III : ORF1ab- A3209V, V3718A, T3750I, G5063S, P5401L and Spike-A222V; Sub-lineage –IV ORF1ab- P309L, D2980N, F3138S and spike - K77T. This study indicated that majority of the clinical cases in the breakthrough were infected with the Delta variant and only 9.8% cases required hospitalization while fatality was observed in only 0.4% cases. This clearly suggests that the vaccination does provide reduction in hospital admission and mortality.

## Introduction

Severe acute respiratory syndrome coronavirus-2 (SARS-CoV-2) was reported from Wuhan in December 2019 and rapidly spread across the globe. The World Health Organization (WHO) declared it as a Public Health Emergency of International Concern on 11th March, 2020. Since then, the virus is continuously evolving and the first major mutation was seen in Spike-protein (D614G) which led to increased infectivity[1]. However, several new SARS-CoV-2 variants of concern (VOC) i.e., Alpha (B.1.1.7), Beta (B.1.35) and Gamma (B.1.1.28.1) have been detected from United Kingdom, South Africa and Brazil respectively during September to December 2020 and have also been reported from India[2–4]. Our earlier study of genomic surveillance from January-August 2020 showed absence of VOC/VUI and presence of G, GR and GH clade in the country with number of mutations[5]. The circulation of variants globally, enhanced the effect of COVID-19 pandemic with increased transmissibility, enhanced severity of illness, diminished protection relative to previous SARS-CoV-2 variant infection, lower response to vaccines and monoclonal antibodies[6–8].

Since the worldwide alert of VOCs, the international travellers arriving at Indian airports from different countries from December 2020 till date were tracked and subjected to SARS-CoV-2 specific real time RT-PCR. The genomic surveillance led to the detection of VOCs i.e., Alpha and Beta; variant of interests (VOIs) i.e., Eta (B.1.525), Kappa (B.1.617.1) and Zeta (B.1.1.28.2) variant under monitoring i.e., B.1.617.3[2–5,9,10]. The recent emergence of the B.1.617 lineage has created a grave public health problem in India. The lineage evolved further to generate sub-lineages B.1.617.1, B.1.617.2, B.1.617.3[11]. Apparently, the sub-lineage B.1.617.2 has gradually dominated the other variants including B.1.617.1, B.617.3 and Alpha VOC in Maharashtra state [9,10]. This variant has further evolved into two new strains called as Delta AY.1 and Delta AY.2.

Several candidate vaccines have been developed using various platforms on fast track mode. Many of them have been used in different countries across the globe under emergency use authorization (EUA). On January 2, 2021 the National Regulatory Authority of India accorded restricted emergency use authorization to the viral vector vaccine developed by Oxford– AstraZeneca (Covishield, manufactured in India) and inactivated vaccine, BBV152 (Covaxin). Subsequently Sputnik V received EUA on 13^th^ April, 2021. In India, National COVID-19 vaccination program was launched on 16^th^ January, 2021 wherein 34,78,36,387 people given vaccine of 28,47,84,116 received two doses of vaccine while 6,30,52,27 persons have got one dose of vaccine as of 6^th^ July 2021[12].

Protection offered by vaccines is being questioned following the emergence of VOCs and reduced real world effectiveness of certain candidate vaccines against these variants. Israel reported breakthrough COVID-19 infections in individuals immunized with the Pfizer vaccine on 9^th^ April, 2021[13]. India started observing a second upsurge in the number of COVID-19 cases from March 2021 onwards followed by a devastating second wave. Further in April, 2021 reduction in neutralization capacity of Covishield/AstraZeneca vaccinated sera against the B.1.1.7 variant as compared to the ancestral strain was observed during *in-vitro* studies[14,15]. Following this, in mid-April, 2021 we decided to trace breakthrough COVID-19 infections across the country and gain cognizance of the different variants which were responsible for the infections.

During April 2021, Hacisuleyman et al., reported 417 cases of breakthrough infections in individuals vaccinated with Pfizer and Moderna mRNA vaccines[16]. Further, post vaccination breakthrough COVID-19 infections are being reported from all across the globe. Centre for Disease Prevention and Control (CDC), reported a total of 10,262 COVID-19 vaccine breakthrough infections[17]. Breakthrough infections in healthcare workers vaccinated with the BNT162b2 vaccine were reported from Italy in the month of May, 2021 during an outbreak of SARS-CoV-2 lineage B.1.1.7[18]. In India, few studies reported breakthrough infections in small parts of the country like Kerala[19] and Delhi[20]. Taking cognizance of such reports, in April-May, 2021, a nationwide study was undertaken to understand the clinico-demographic profile of patients and SARS-CoV-2 strains responsible for post vaccination breakthrough COVID-19 infections across the country. To our understanding, this is the largest and first nation-wide study of post-vaccination breakthrough infections from India.

## Materials and Methods

### Study Catchment area

The Indian Council of Medical Research -Department of Health Research (ICMR-DHR) utilized network of Viral Research and Diagnostic Laboratories (VRDLs) to track breakthrough infections. breakthrough cases defined as the detection of SARS-CoV-2 RNA or antigen in a respiratory specimen collected from a person ≥14 days after receipt of all recommended doses of an COVID-19 vaccine. Clinical and demographic details as well as nasopharyngeal/oropharyngeal swabs (NPS/OPS) of SARS-CoV-2 patients (n=677) tested positive by real-time RT-PCR were collected by the VRDLs in the North, South, West, East, North-East, and Central parts of India from 17 states and Union Territories (UTs) (Maharashtra, Kerala, Gujarat, Uttarakhand, Karnataka, Manipur, Assam, Jammu and Kashmir, Chandigarh, Rajasthan, Madhya Pradesh, Punjab, Pondicherry, New Delhi, West Bengal, Tamil Nadu, New-Delhi) during April-May, 2021. These clinical specimens were sequenced using next generation sequencing (NGS) to determine nucleotide variations in the SARS-CoV-2 genome from these strains.

### Inclusion, exclusion criteria and transport of specimens

Cases fulfilling the following inclusion criteria were enrolled under the study: i) cases who contracted COVID-19 infection after the second dose of COVID-19 vaccine (Covaxin or Covishield) with or without previous history of COVID-19 ii) Cases whose real time RT-PCR threshold value was <30 and NPS/OPS were appropriately stored at -80 °C; (iii) Sample referral forms (SRF) capturing the demographic and clinical details of cases were available with the respective VRDLs. All the specimens of breakthrough cases fulfilling the above criteria were packed in triple-layer packaging with dry ice as per International Air Transport Association (IATA) guidelines and then transferred to the Reference laboratory the ICMR-National Institute of Virology (ICMR-NIV), Pune for sequencing and variant analysis. In April 2021, a new specimen referral form (SRF) which included details of COVID-19 vaccination was launched by ICMR throughout the country. However, quite a few states had not implemented this new SRF. Therefore, it was not possible to track COVID-19 breakthrough infections in these states and they were not included in this study.

### Retrieval of clinical and demographic data

Though completely filled SRFs were requested along with the specimens, most of the forms received from laboratories were incomplete due to increased testing load during the second wave of COVID-19 in India. Therefore, telephonic interviews were conducted wherein each breakthrough case was called and interviewed individually during the period of 25th May to 14th June, 2021. The telephonic interviews also helped in validating the information available in the SRF and to fill in missing data. The patients were questioned on their demographic details, vaccination details, history of earlier COVID-19 infection, history of contact with laboratory confirmed case of COVID-19 cases prior to breakthrough infection, presence of co-morbidities, symptoms developed and course of infection including the details of hospitalization. Each phone call typically lasted for 10-12 minutes and only patients who elicited a complete history were included in the study. A total of 814 clinical specimens were received from the different VRDLs all over the country at ICMR-NIV, Pune. The onset date/OPS and NPS collection dates ranged from 5th March to 3rd June, 2021, which coincided with the second wave of COVID-19 pandemic in India. Out of these, 15 patients were not vaccinated for COVID-19, while 2 patients didn’t give any vaccination history, and 120 patients could not be traced. Thus, a total of 137 patients were excluded and a total of 677 cases were included in the study.

### RNA extraction and Next generation sequencing

Total RNA was extracted from 200-400 μl of NS/OS swab samples using an automated RNA extraction system (Thermofisher, USA) using Magmax RNA extraction kit (Applied Biosystems, USA). Real time RTPCR was set using SARS-CoV-2 specific primers for the detection of E gene and RdRP gene as described earlier[21]. Illumina Covidseq protocol (Illumina Inc, USA) was followed for preparation of RNA libraries. Extracted RNA was annealed using random hexamers to prepare for cDNA synthesis. First strand cDNA was synthesized using reverse transcriptase. The synthesized cDNA was amplified in two separate PCR plates using two pools of primers (Pool 1 and Pool 2) covering entire genome of the SARS-CoV-2. Amplified cDNA was then tagmented and bead-based post tagmentation clean-up was performed. Tagmented amplicon were further amplified in this step using a PCR program as per manufactures instructions (Covidseq reference guide, Illumina). This PCR step adds pre-paired 10 base pair Indexes (Set 1, 2, 3, 4 adapters), required for sequencing cluster generation. One Covidseq positive control (CPC) and one negative template control (NTC) were used for each 96-well plate. Libraries generated in batch of 96 samples per plate were pooled into one 1.7 ml tube. Libraries of optimal size were purified by using magnetic bead based cleanup process method. Amplified and purified libraries were quantified using KAPA Library Quantification Kit (Kapa Biosystems, Roche Diagnostics Corporation, USA).

For a set of 384 samples, combined 25 µl of each normalized pool containing index adapter set 1, 2, 3, 4 in a new micro-centrifuge tube. At this step a final pool of 384 samples were diluted to a starting concentration of 4 nM. These libraries were then denatured, diluted and then loaded at final loading concentration of 1.4pM on to the NextSeq 500/550 system using NextSeq 500/550 High Output Kit v2.5 (75 Cycles) as per the manufacturer’s instructions (Illumina Inc., USA). The files were analyzed using the reference-based mapping method as implemented in CLC genomics workbench version 20.0 (CLC, QIAGEN, and Germany). The Wuhan Hu-1 isolate (Accession Number: NC_045512.2) was used as the reference sequence to retrieve the genomic sequence of the SARS-CoV-2. The retrieved sequences were aligned along with few representative sequences from GISAID database in CLC genomic Work bench v.20. A phylogenetic tree was generated using the MEGA software [22] for the aligned sequences. Gene wise amino acid mutations were also observed.

## Results

### Clinical and demographic analysis of the breakthrough samples

Detailed distribution of breakthrough cases (n=677) collected from 17 states/UT of the country used for NGS is provided in Table-1. The clinical samples for the analysis were collected between March-June, 2021. Out of these 677 patients, 85 acquired COVID-19 infection after taking first dose of the vaccine, while 592 were infected after receiving both doses of vaccine. Similarly, the clinical samples from the COVID-19 cases post second dose of vaccination were collected with a median of 38 days and had an inter-quartile range of 20 (19-58) days. A total of 604 patients had received Covishield vaccine, 71 had received Covaxin and two had received Sinopharm vaccine (BBIBP-CorV).

**Table 1:** Region-wise and state-wise distribution of SARS-CoV-2 clinical samples used for Next-generation sequencing (n=677):

| **Region** | **State/UTs** | **Clinical Samples received from each site** |
| --- | --- | --- |
| North India | New-Delhi | 20 |
|  | Uttarakhand | 50 |
|  | Jammu & Srinagar | 25 |
|  | Punjab | 12 |
|  | Chandigarh | 19 |
| North-Eastern India | Manipur | 15 |
|  | Assam | 40 |
| Eastern India | West Bengal | 10 |
|  | Jharkhand | 12 |
| Central India | Madhya Pradesh | 68 |
| Western India | Maharashtra | 53 |
|  | Gujarat | 47 |
|  | Rajasthan | 58 |
| South India | Karnataka | 181 |
|  | Kerala | 16 |
|  | Tamil Nadu | 25 |
|  | Puducherry | 26 |

Clinical data was analyzed for 677 breakthrough cases. The median age (and the IQR) of patients in the study was 44 (31-56), with the breakthrough cases after one dose was 53 (45-61) and after two doses was 41(30-55). A total of 441 (65.1%) of the breakthrough cases were males. A total of 482 cases (71%) were symptomatic with one or more symptoms, while 29% had asymptomatic SARS-CoV-2 infection. Fever (69%) was the most consistent presentation followed by body ache including headache and nausea (56%), cough (45%), sore throat (37%), loss of smell and taste (22%), diarrhoea (6%), breathlessness (6%) and 1% had ocular irritation and redness. The clinical and demographic analysis of the 677 cases of breakthrough infections are is enumerated in Table 2.

**Table 2:**
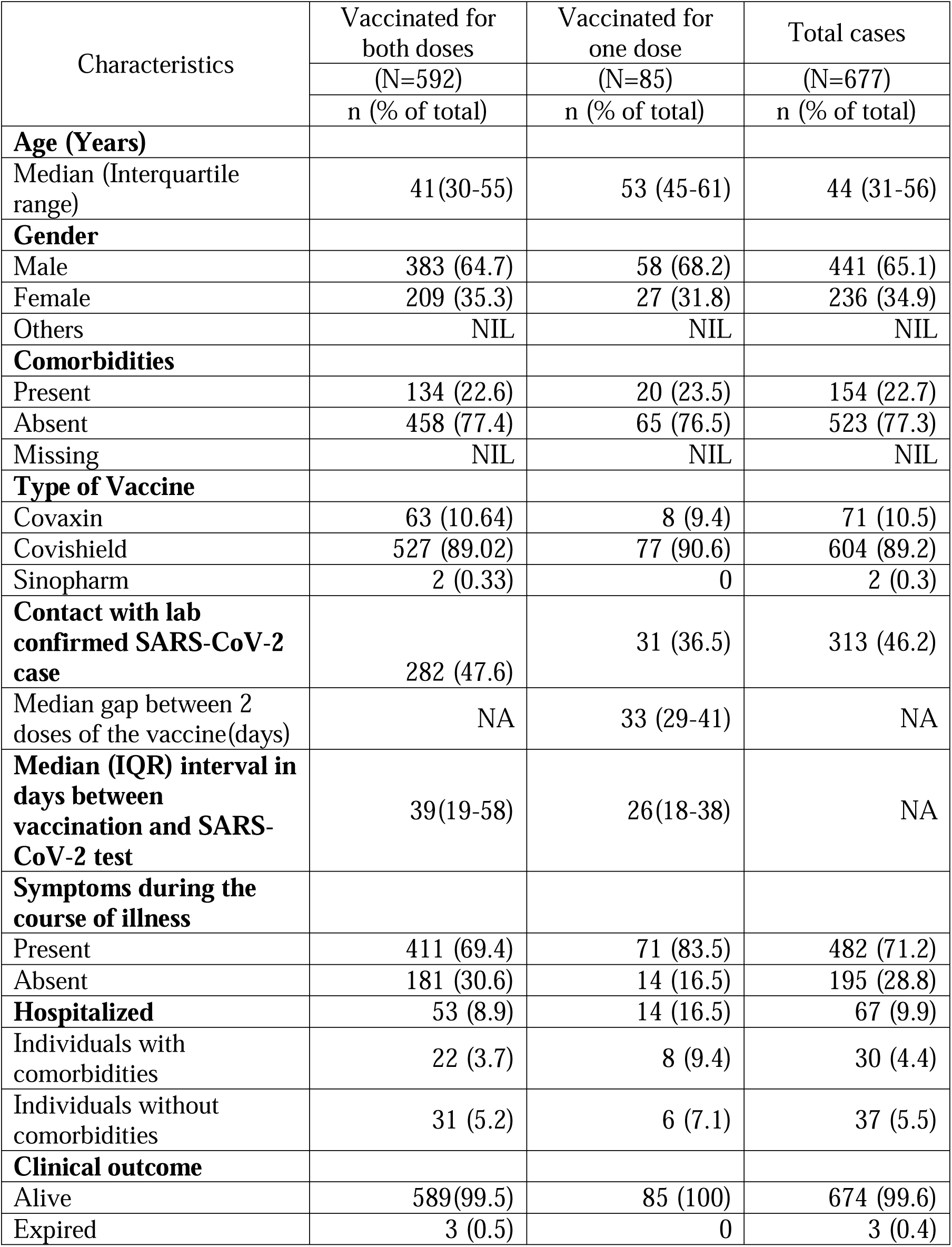
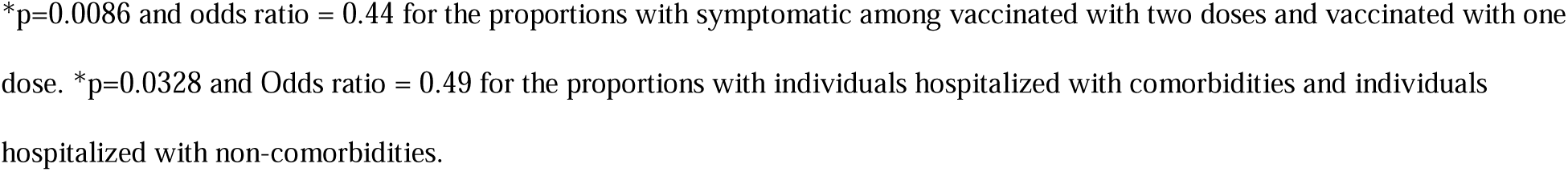
Demographic analysis of breakthrough COVID-19 infections

Co-morbidities were observed in the 154 out of 677 cases which included diabetes mellitus type-II, hypertension as well as chronic cardiac, renal and pulmonary diseases and obesity. The symptoms reported in patients with breakthrough infections are enumerated in Figure 1. The co-morbid cases were significantly pre-disposed to develop symptoms [cough, sore throat, fever, loss of smell and taste, diarrhoea, breathlessness, ocular symptoms and constitutional symptoms (body ache, headache, nausea)]; (OR=2.0042, 95% C.I.=1.2857 to 3.1244, z-statistic=3.069, *p=0*.*0021*). Also the co-morbid breakthrough cases were significantly more predisposed to hospitalization (OR=3.1779, 95% C.I.=1.8886 to 5.3471, z-statistic=4.355, *p<0*.*0001*).

**Figure 1:**
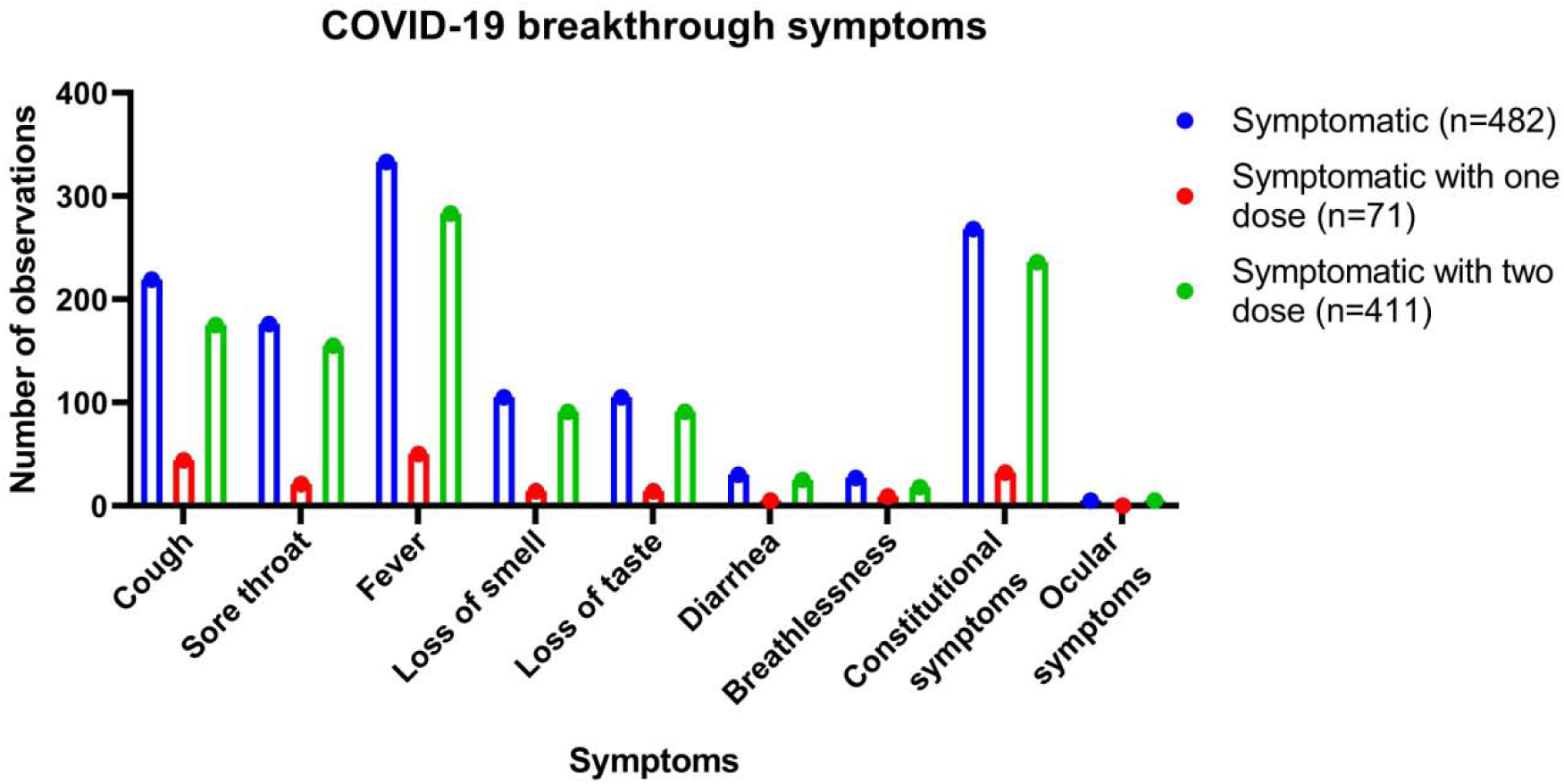
Symptoms reported in COVID-19 breakthrough infections

### Next-generation sequencing analysis of the breakthrough specimens

Out of 677 cases included in this study, 593 were the clinical sample from breakthrough and 84 had infection after one dose. Sequencing was not performed for 112 clinical samples (break through: n=95; single dose: n=17) based on the higher Ct and less Kappa value.

The complete genome of 511 SARS-CoV-2 was recovered with genome coverage of more than 98% [break through: n=446; single dose: n=65]. SARS-CoV-2 sequences with more than 99 and 84% of the genome coverage were recovered from 446 [breakthrough: n=387; single dose: n=59] and 546 [break through: n=480; single dose: n=66] clinical specimens respectively. Less than 98% genomes were retrieved from 54 samples and were not used further in analysis. The details of the percentage genome retrieved, total reads mapped and percent relevant reads are given in Supplementary Table 1. The lineages were retrieved using the Pangolin online software (https://cov-lineages.org/pangolin.html) from the specimens with more than 84% genome coverage and mentioned in Supplementary Table 1.

The distribution of the different SARS-CoV-2 variants with 98% genome coverage were characterised using Pangolin software and presented in Figure 2 of the studies area. It is observed that Delta [B.1.617.2] (n=384) was the major SARS-CoV-2 lineage observed in the breakthrough samples followed by B.1.1.7 (n=28). Kappa [B.1.617.1] (n=22), B.1.617.3 (n=2), B (n=1), B.1.36 (n=5), B.1.1.294 (n=1), B.1.36.16 (n=1), B.1.1.306 (n=1), and Delta AY.2 (n=1) pangolin lineages variants were also observed along with others details are given in Supplementary Table 1. Sixty-five samples out of 84 samples from individuals infected with SARS-CoV-2 after one dose infection had 99.5% genome retrieval. These sequences had Delta [B.1.617.2] (n=59), B.1.1.7 (n=4) Kappa [B.1.617.1] (n=1), Delta AY.1 (n=1). The Delta AY.1 variant was observed in Madhya Pradesh (MP) while Delta AY.2 was observed in the Rajasthan state of India. The percent nucleotide divergence of the different SARS-CoV-2 strains relative to reference was 99.81-100% details for each strain are given in Supplementary Table 1.

**Figure 2.**
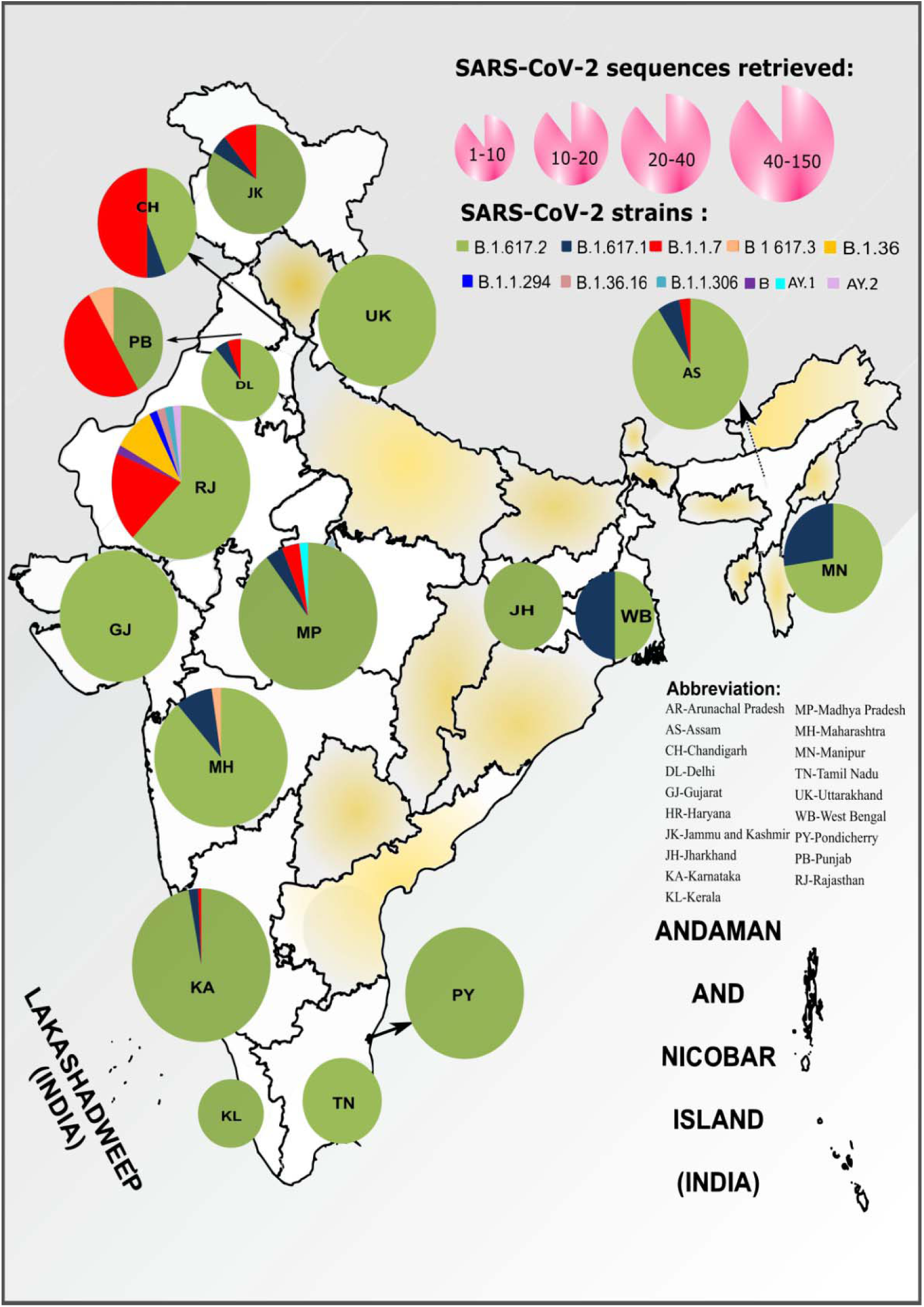
Distribution of the SARS-CoV-2 genome prevalence in the breakthrough phase (March-June 2021) during the second wave of SARS-Cov-2 infection: The size of each pie chart within the states of the Indian map is ranged based on the number of sequences retrieved in the study. The distribution in the pie chart is proportional to the numbers in each respective clade in each state. The outline of India’s map is downloaded from http://www.surveyofindia.gov.in/file/Map%20of%20India_1.jpg (accessed on 20 March 2020) and further modifie to include relevant data in the SVG editor

It was observed that Southern, Western, Eastern and North-Western regions of India predominantly reported breakthrough infections from mainly Delta and then Kappa variant of SARS-CoV-2. The Northern and central regions reported such infections due to Alpha, Delta and Kappa variants; however, cases due to Alpha variant predominated in the northern region (Figure 2). The overall majority (86.09%) of the breakthrough infections were caused by the Delta variant (B.1.617.2) of SARS-CoV-2 in different regions of India except for the northern region where the Alpha variant predominated.

Figure 3 depicts the neighbour-joining tree generated using the Tamura-3-parameter model with a bootstrap replication of 1000 cycles. SARS-CoV-2 sequences (n=421) with genome coverage of 99% and fewer gaps in coding regions were taken for the generation of a phylogenetic tree. Thirty–two representative and 421 SARS-CoV-2 sequences retrieved in this study were used to generate the phylogenetic tree. The Delta sequences (n=358) presented the highest representation of breakthrough cases from different parts of the country and clustered into 4 distinct sub-lineages. Sub-lineage-I had 166 number of SARS-CoV-2 sequences, while sub-lineages-II, -III and –IV had 100, 68, and 24 sequences respectively and are marked in the phylogenetic tree. The gene-wise amino acid mutations were further looked upon for the retrieved sequences and the representative sequences relative to the reference sequence. It was observed that the Delta SARS-CoV-2 variant sequences had conservation in different gene positions leading to differential clustering. These conserved mutations of different sub-lineages are depicted in Figure 4. **Sub-lineage –I (red colour):** ORF1ab-A1306S, P2046L, P2287S, V2930L, T3255I, T3446A, G5063S, P5401L, A6319V and N-G215C; **Sub-lineage –II (green colour):** ORF1ab-P309L, A3209V, V3718A, G5063S, P5401L and ORF7a-L116F; **Sub-lineage –III (pink colour):** ORF1ab-A3209V, V3718A, T3750I, G5063S, P5401L and Spike-A222V; **Sub-lineage –IV (Orange colour):** ORF1ab-P309L, D2980N, F3138S and spike - K77T. **Common in B.1.617.2 lineage**: ORF1ab-P4715L, spike - T19R, L452R, T478K, D614G, P681R, ORF3a-S26L, M-I82T, ORF7a-V82A, T120I and N-D63G, R203M, D377Y.

**Figure 3.**
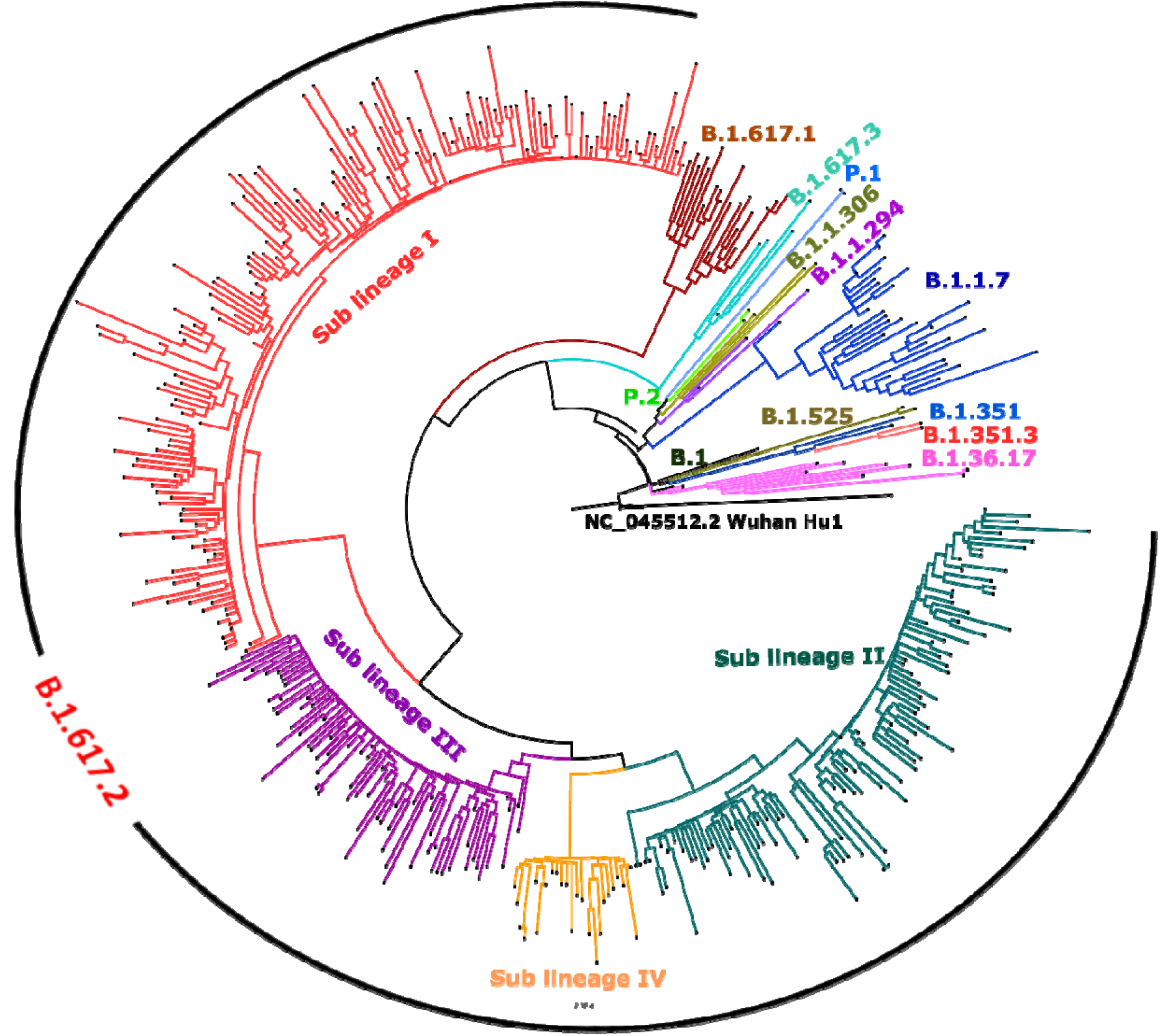
Phylogenetic tree of the 402 SARS-CoV-2 genomes from breakthrough: A Neighbor-joining tree of the 402 SARS-CoV-2 sequences retrieved in this study, along with the representative SARS-Cov-2 sequences from different clades with a bootstrap replication of 1000 cycles. Four major sub-lineages of delta variant were observed, which are marked on branched in different colours. Sub-lineage-I-IV is marked in red, green, pink and orange colour on the nodes respectively. B.1.617.1 sequence is marked in brown and B.1.617.3 in blue colour. Th representative pangolin lineages are also marked on branches in different colours. FigTree v1.4.4 and Inkscap were used to visualize and edit the generated tree.

**Figure 4.**
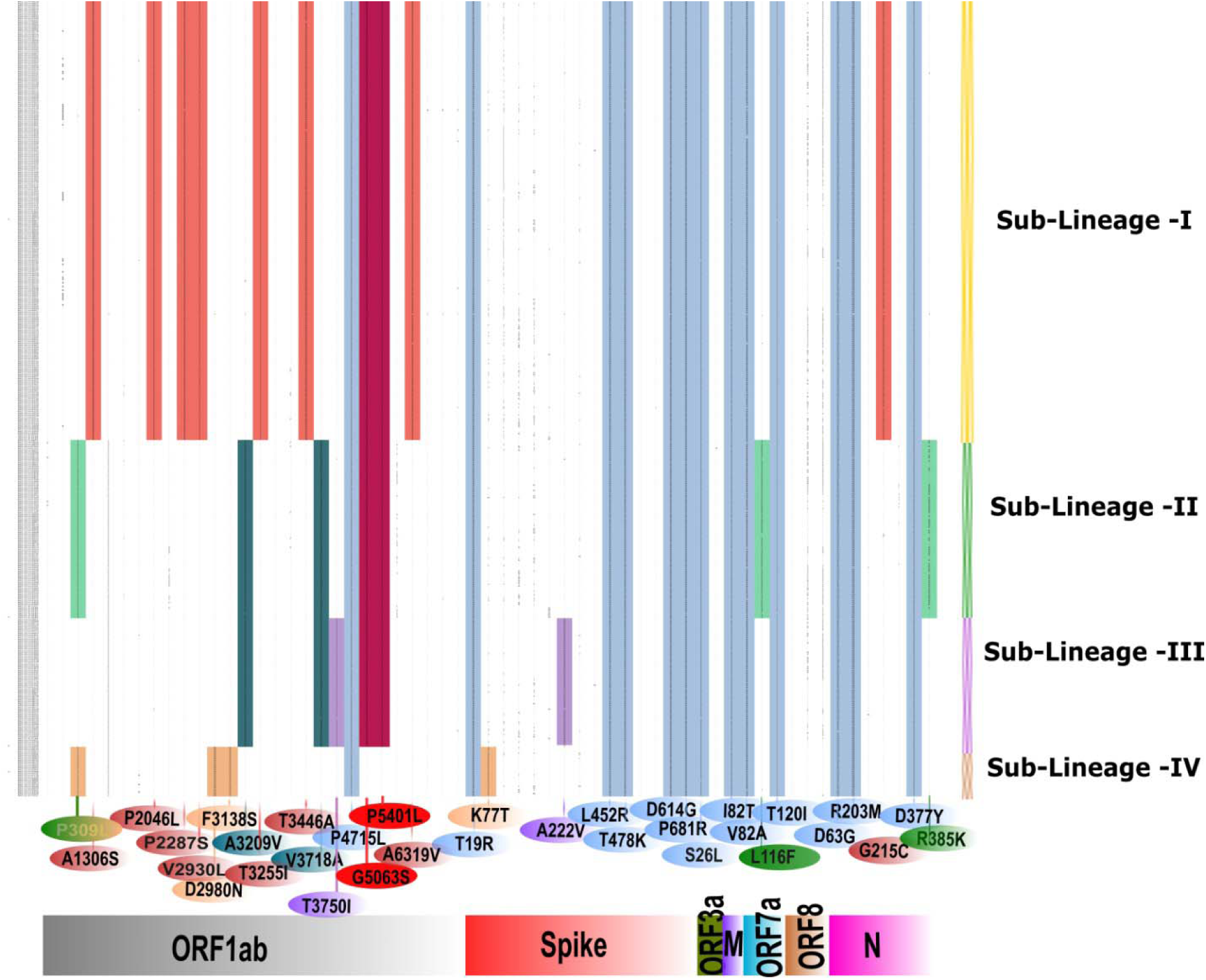
Characterization of sub-lineages observed in the delta SARS-CoV-2 variant from breakthrough sequences: Sub-lineage-I-IV is marked in red, green pink and orange colour along Y-axis. The amino acid mutations observed in different genes are marked on the X-axis. The amino acid change is marked concerning Wuhan-HU-1 (Accession No: NC_045512.2) It is observed that a couple of mutations are conserved in sub-lineages I-III and marked as the gradient of red-violet colour. Amino acid mutations conserved for sub-lineage II and III are marked as a violet-green gradient. The amino acid changes common to Delta variant is marked in blue colour

## Discussion

Globally, COVID-19 vaccines were accorded EUA and introduced quickly into the public health program to prevent SARS-CoV-2 infections and curtail disease transmission, thus saving lives and livelihood. However, the emergence of SARS-CoV-2 VOCs has raised a public health concern due to increased transmissibility and potential to evade humoral immune response. Apparently, it has also coincided with the introduction of global COVID-19 vaccination program which has created dilemma about the efficacy of vaccines under EUA. Several studies have reported the reduction in the neutralization potential of the sera of individuals vaccinated with ChAdOx1 nCoV-19 (86×), mRNA-1273 (8.6×), BNT162b2 (6.5×) and BBIBP-CorV (1.6×) against Beta variant[23]. Beside this, the vaccine sera of BNT162b2 mRNA (7×) and one dose of ChAdOx1 nCoV-19 had also shown reduced neutralization against Delta variant[24].

As per the WHO classification, the Delta variant has been designated as a variant of concern due to increased transmission and higher immune evasion, whereas other two sub-lineages of B.1.617 namely B.1.617.1 and B.1.617.3 with E484Q are grouped in VUI[25]. B.1.617 variant and its lineage B.1.617.2 were primarily responsible for the surge in COVID-19 cases in Maharashtra state[26]. Delta (B.1.617.2) and Kappa (B.1.617.1) was detected among 60% of the clinical specimens of the COVID-19 cases collected from Maharashtra during January and February 2021[26]. The rapid rise in the daily infection was observed in India with dominance of Delta variant which accounted for >99% of all sequenced genomes in April 2021[27]. A recent study on the secondary attack rates in UK households demonstrated a higher transmission of Delta compared to the Alpha variant[8]. A reduced neutralizing capability of currently used SARS-CoV-2 vaccines against Delta variants which is one of the causes for recent increase in the breakthrough cases with this strain.

Emergence of variant of concerns has led to upsurge in COVID-19 cases and second wave of pandemic in various countries including India. Incidentally, several countries have reported COVID-19 breakthrough infections even after completion of full vaccination schedule[16,18,28]. More than 10,000 breakthrough infections after completion of full course of vaccination has been reported in USA. Overall breakthrough infections were seen in smaller percentage of total vaccinated population[17]. A recent study has also reported mild symptomatic breakthrough infections from Kerala and Delhi, India[19,20].

The present study revealed the infection among breakthrough cases predominantly occurred through Delta variant indicating its high community transmission during this period followed by Alpha and Kappa variants. In our study, 67 cases (9.8%) required hospitalization and fatality was observed in only 3 cases (0.4%). This clearly suggests that vaccination reduces severity of disease, hospitalization and mortality. Therefore, enhancing the vaccination drive and immunizing the populations quickly would be the most important strategy to prevent further deadly waves of the COVID-19 and would reduce the burden on the health care system.

Two new SARS-CoV-2 variants Delta AY.1 and AY.2 were also identified in these study samples. Delta AY.1 and AY.2 is characterized by the presence of K417N mutation in the spike protein region. K417N, E484K, L452R, and E484Q are the mutations known to disrupt receptor binding domain (RBD) binding capacity and making them more infectious by immune escape mechanism against the current vaccines[29]. This indicates improved virus fitness to evade immune responses and survive against the vaccines.

Post-vaccination breakthrough COVID-19 infections have been reported from various countries with use of different licensed vaccines. It appears that the current COVID-19 vaccines are disease modifying in nature wherein mild or less severe infections are expected to occur in vaccinated individuals. However, vaccination seems to have an obvious advantage in averting severe disease, hospitalization and deaths. Therefore, continuous monitoring of post vaccination breakthrough infections along with clinical severity of disease must be adopted as an essential component of vaccine roll-out program by all countries. Such monitoring will help us to understand the need to adequately tweak the available vaccines and also develop new vaccines with enhanced potential to protect against variant strains of SARS-CoV-2.

Identification of the new variants that is responsible for the breakthrough infections underline the importance of this study. It also highlights the need of active genomic surveillance of the new SARS-CoV-2 variants and assessing their potential to evade the immune response.

## Ethical approval

The study was approved by the Institutional Human Ethics Committee of ICMR-NIV, Pune, India under project ‘Molecular epidemiological analysis of SARS-CoV-2 circulating in different regions of India’.

## Data Availability

All the data form this study is available with the corresponding author on request.

## Acknowledgement

**A**uthors gratefully acknowledge the encouragement and support extended by Prof. (Dr) Balram Bhargava, Secretary to the Government of India Department of Health Research, Ministry of Health & Family Welfare & Director-General, ICMR and Dr. Samiran Panda, ECD Chief, ICMR, Delhi. We thank the team member of Maximum Containment Facility, ICMR-NIV, Pune including Dr. Deepak Patil, Ms, Pranita Gawande, Ms. Kaumudi Kalele, Mrs. Ashwini Waghmare, Ms. Tejashri Kore, Ms. Shilpa Ray, Ms. Priyanka Waghmare and Ms. Poonam Bodke for excellent support. The authors would like to acknowledge Krishnapal Karmodia and Sanjeev Galande from IISER, Pune for helping us utilize their NGS facility. Further, we also acknowledge Anjani Gopal from IISER, Pune for assisting us in the NGS facility. We would like to acknowledge all the authors that have submitted the SARS-CoV-2 sequences to the GISAID database.

## Author Contributions

Conceptualization, P.D.Y., N.G.; methodology, P.D.Y., N.G. H K, L.M, R.R.S, A.K, D.A.N, A.M.S, S.P, T.M, M.D, P.P, Y.J; software, D.A.N, A.K, M.D, P.P, Y.J.; re-sources, S.R, S.G, J.N, N.V, N.A, G.N, A.K.B, M.P.K, D.B, P.B, J.I, S.R, S.D, S.D, P.B, P.G, B.B, D.K, K.D, B.F, K.G, R.J, A, R.D, S.D, D.K, N.K, Y.K.T, P.K.K, A.G, H.K, B.M, M.N, L.D, N.S,; writing original draft preparation, P.D.Y., N.G., D.A.N.; writing, P.D.Y., N.G., H.K, D.A.N supervision, P.D.Y., N.G., R.S.S, A.M.S.; project administration, P.D.Y., N.G., P.A.; funding acquisition, P.D.Y., P.A. All authors have read and agreed to the published version of the manuscript.

